# Encapsulated Salmon Polar Lipids Modestly Reduce Postprandial Platelet Sensitivity to PAF and Thrombin in Healthy Adults: A Pilot Study

**DOI:** 10.64898/2025.12.02.25341455

**Authors:** Ronan Lordan, Alexandros Tsoupras, Phil Jakeman, Ioannis Zabetakis

## Abstract

The postprandial effects of a novel food-grade extracted salmon polar lipids (SPL) supplement against platelet-activating factor (PAF) and thrombin-induced platelet aggregation in a human pilot study was evaluated. This study was double-blinded, crossover, and placebo controlled in design. Five healthy volunteers completed 4 h time-course trials on 5 separate days. Blood was drawn at baseline before subjects consumed a standardized breakfast with one of the following treatments: a low-dose (0.25 g) or a high-dose (0.5 g) of SPL encapsulated within a stomach resistant capsule (LDSR and HDSR respectively) or a non-resistant capsule (LDSNR and HDSNR, respectively), or placebo capsules containing food-grade glycerin (0.5 g; Placebo). Blood was analyzed at 1 h intervals for 4 h. Among the treatments tested, the high-dose non-resistant capsule (HDSNR) produced the clearest postprandial effects, inducing significant increases in EC_50_ (reduced platelet sensitivity) for PAF at 2–3 h and for thrombin at 3–4 h post-ingestion. Smaller or delayed effects were observed with the high-dose resistant capsule (HDSR), while low-dose formulations produced minimal changes. Postprandial plasma glucose, lipid profile (TC, HDL-C, LDL-C, TG), fibrinogen, prothrombin time, and activated partial thromboplastin time remained unaffected across all trials. This pilot study provides the first in vivo indication that SPL supplementation may modestly attenuate platelet responsiveness to both PAF and thrombin without altering standard haemostatic or metabolic biomarkers. Larger controlled studies are warranted to confirm these preliminary findings and define optimal dosing and formulation strategies.

## 1. Introduction

Low-grade or systemic inflammation contributes to the onset of atherosclerosis and subsequent cardiovascular diseases (CVD) [1–4]. Activation and aggregation of platelets contribute to the ‘crosstalk’ between various cells including endothelial cells and leukocytes engaged in the inflammatory development of atherosclerosis and atherothrombotic events [5–7]. Platelet agonists mediate platelet activation and aggregation. Of these, platelet-activating factor (PAF) and thrombin are among the most potent [5,8,9]. PAF activates various pathophysiological signaling pathways in response to inflammatory stimuli that negatively affect cardiovascular risk factors [10,11]. Thrombin on the other hand, is a procoagulant and pro-inflammatory serine protease that participates in coagulant catalytic processes as well as the activation of other cell types in several pathophysiological processes [8,9,12], including atherosclerosis [12]. PAF and thrombin are known to enhance the expression of cell adhesion molecules that induce secretion of pro-inflammatory cytokines, an inflammatory response in atherosclerotic plaques, proliferation of aortic smooth muscle cells and vascular lesion at sites of injury [2,12,13].

The inhibition of PAF and thrombin related pathways offers a potential therapeutic strategy to target thrombosis and inflammatory responses that underpin the atherosclerosis and inflammatory disorders [2,13,14]. It is recognized that some dietary patterns and their components have the potential to reduce levels of specific risk factors for CVD, including the reduction of inflammation [15,16]. CVD-related PAF and thrombin pathways are affected by the diet either favorably, by positive dietary behaviors such as those attributed to the Mediterranean diet, or detrimentally by dietary behavior related to the Western diet [17]. The Western diet is characterized by energy-rich, ultra-processed foods that cause exaggerated postprandial increases in plasma lipids and glucose leading to hyperglycemia and hyperlipidemia [18]. These elevations lead to excess pro-inflammatory reactions and activated platelets and immune cells [19,20]. Indeed, postprandial platelet activation and hypercoagulability are major risk factors for CVD [21], of which the PAF pathway can be modulated by dietary components postprandially [22].

Fish oils, particularly omega-3 polyunsaturated fatty acids (n-3 PUFA) have been a promising but contentious area of research due to their putative cardioprotective effects [23,24]. Food-grade extracts from sustainable sources are potential candidates for the development of novel cardioprotective nutraceuticals, which are important for improving underlying health conditions and overall well-being in aging populations characterized by increased risk of non-communicable diseases [25,26]. In previous studies, it has been demonstrated that polar lipids extracted from Irish organic farmed salmon (*Salmo salar*) possess strong antithrombotic properties against platelet aggregation *in vitro* relating to the PAF and thrombin pathways *in vitro* [25,27,28]. Studies have also shown that salmon oil may favorably affect metabolic syndrome through phospholipid remodeling, inhibiting insulin resistance, and reducing inflammatory factors [29]. Indeed, the consumption of fish and fish oil supplements may affect platelet function and cardiovascular health [30–32], although there is a dearth of postprandial investigations assessing the biological effects of marine-sourced polar lipids. This pilot study investigates the temporal pattern and magnitude of the antiplatelet effects of food-grade extracted salmon polar lipid ingestion (SPL) postprandially in a pilot study of five healthy human subjects over a 4 h period.

## 2. Materials and Methods

### 2.1. Materials and instrumentation

All glass and plastic consumables, reagents, and solvents were of analytical grade and were purchased from Fisher Scientific Ltd (Dublin, Ireland). 18G Insyte™ cannulas were purchased from Becton Dickinson Ltd. (UK). A CARESITE® extension set, 0.9% w/v saline, and Omnifix® syringes with varied volumes were purchased from Braun Medical Ltd. (Dublin, Ireland). Tegaderm™ transparent protective dressings were acquired from 3M Health Care (Neuss, Germany). Evacuated sodium citrate 9 NC, EDTA KE, glucose FE, and serum gel Z S-monovettes for blood sampling were purchased from Sarstedt Ltd. (Wexford, Ireland). The *ex vivo* platelet aggregation bioassay was carried out on a Chronolog-490 two channel turbidimetric platelet aggregometer (Havertown, PA, USA), coupled to the accompanying AGGRO/LINK software package. All platelet aggregation consumables were purchased from Labmedics LLP (Abingdon on Thames, UK). Standard PAF, thrombin, and BSA were purchased from Sigma Aldrich Ltd. (Wicklow, Ireland). Centrifugations were carried out on an Eppendorf 5702R centrifuge (Eppendorf Ltd Stevenage UK). Spectrophotometric analysis was carried out on a Shimadzu UV-1800 spectrophotometer (Kyoto, Japan).

### 2.2. Isolation and encapsulation of food-grade SPL

A sustainable marine source was chosen for these studies; Irish organic farmed salmon (*Salmo salar*). Fresh salmon fillets were homogenized mechanically by blender and total lipids (TL) extracted and further separated into neutral lipid (NL) and polar lipid fraction (SPL) in accordance with EU legislation for food-grade based extractions of fish oil (consolidated Directive 2009/32/EC: https://eur-lex.europa.eu/legal-content/en/ALL/?uri=CELEX:32009L0032) as previously described [25]. Solvents were evaporated from samples using flash rotary evaporation and lipid samples transferred to small vials and remaining solvents evaporated under a stream of nitrogen. The acquired SPL extracts were weighed and encapsulated under a stream of nitrogen into gelatin-coated capsules of two types: stomach non-resistant capsules (SNR) and stomach resistant capsules (SR). Placebo capsules were prepared containing only glycerin. All capsules were prepared external (Propak Health Ltd., Finglas, Dublin, Ireland) to the university and were tested for microbial safety, heavy metals, pesticides, and various other potentially harmful constituents (Eurofins Food Testing Ireland Ltd., Dungarvan, Co. Waterford, Ireland).

### 2.3. Study population

This study was conducted according to the guidelines laid down in the Declaration of Helsinki, approved by the Ethics Committee of the Faculty of Science and Engineering of the University of Limerick and registered with ClinicalTrials.gov NCT03603769. Following written and informed consent, six participants completed a routine clinical, haematological, biochemical, and anthropometrical screening session with a general practitioner to assess suitability. Exclusion criteria were smoking, following a diet regime or special diet, diagnosis of hypertension, metabolic or endocrine disease, gastrointestinal disorders, blood clotting disorders, dyslipidaemia, or a recent history of medical or surgical events. One volunteer that initially participated and was deemed eligible for the study had to terminate their participation due to personal commitments. A total of five participants (n = 5) completed all aspects of the study. Recruitment, screening, and treatment allocation occurred as outlined in the CONSORT flow diagram (Figure 1). The baseline characteristics of the five participants are provided in Table 1 and Table 2 in the supplementary files. Healthy participants were selected for this pilot study for several reasons, including safety assessment and tolerability of an acute dose of the supplement before testing in a clinical population. Furthermore, this approach allowed for assessment of the capsule under controlled baseline physiology in participants with stable platelet profiles in the absence of medication, which would allow for more clear detection of physiological changes directly attributable to the intervention.

**Fig. 1.**
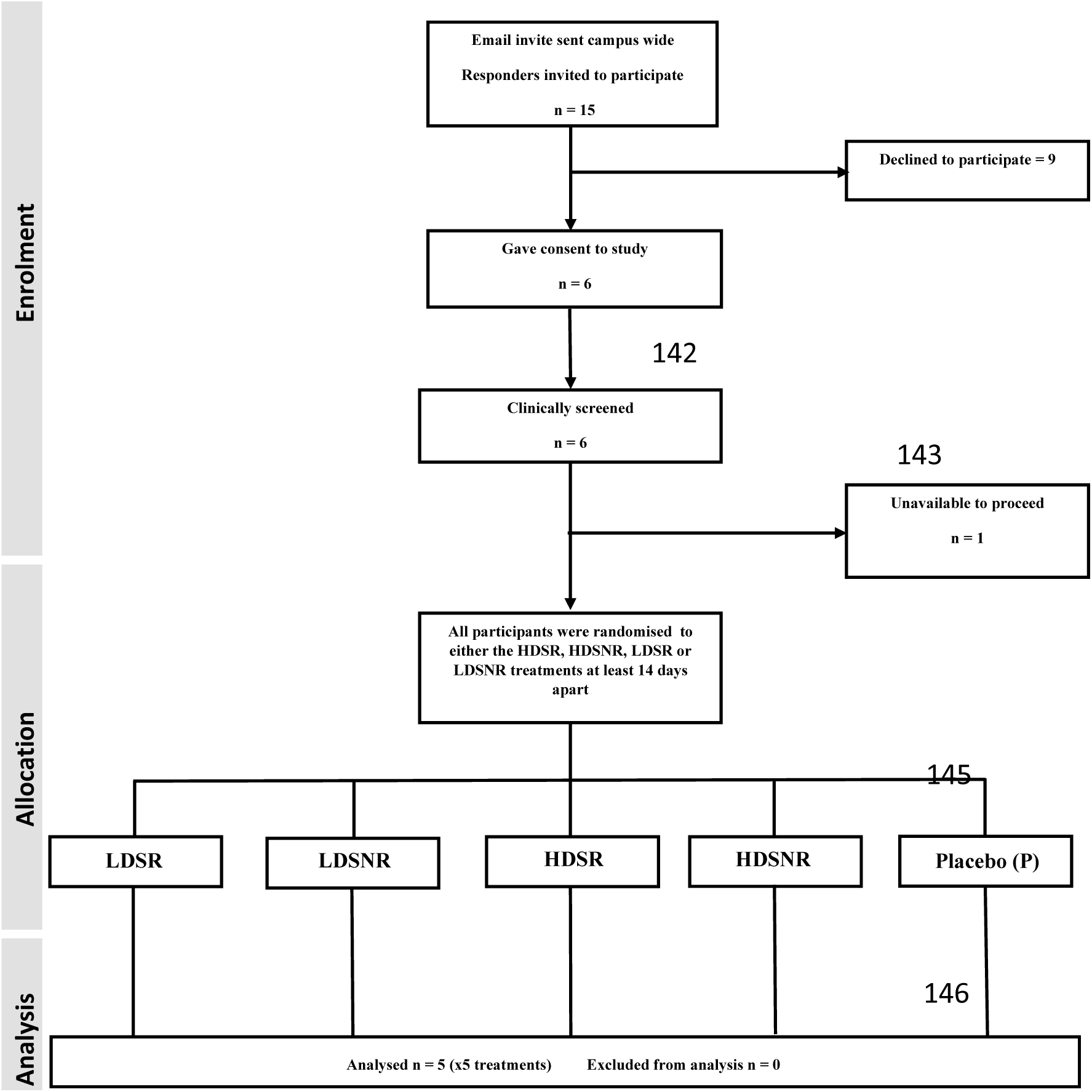
CONSORT flow diagram of participant involvement and trial layout.

### 2.4. Study protocol

In a randomized, double-blind design, each subject completed five trials on separate days at least 14 days apart from each other. Three days before the trial and throughout the entire trial period, volunteers were instructed to follow Irish national dietary guidelines [33], which allows moderate consumption of fish (once or twice per week). The participants were asked to abstain from alcohol, avoid excessive exercise, and sleep at least 7 h/day throughout the study. After fasting overnight (10–12 h), each participant arrived at the laboratory at 09:00 and were seated in a phlebotomy chair and allowed to rest for five minutes. A certified clinical nurse then proceeded to fit the volunteer with a cannula. A 20 G cannula (BD Insyte™) was inserted into the median cubital vein, which was attached to a CARESITE® extension set. The cannula was held in place with a Tegaderm™ transparent protective dressing.

An initial 1.5 mL of blood was drawn through the cannula using a 3 mL syringe. This was discarded to fill the line with blood. A further 21.4 mL of blood was drawn using a 25 mL Omnifix® syringe. The line was then flushed with 10 mL of 0.9% w/v saline using a 10 mL Omnifix® syringe. The collected blood was then transferred into 5 S-monovettes: 1) 8.2 mL sodium citrate 9 NC S-monovette; 2) 2.9 mL sodium citrate 9 NC S-monovette; 3) 2.7 mL EDTA KE S-monovette; 4) 2.7 mL Glucose FE S-monovette; 5) 4.9 mL Serum Gel Z S-monovette (Sarstedt Ltd., Wexford, Ireland). All monovettes were appropriately labelled and placed in clinical sample transportation boxes in order to be sent to the appropriate laboratory for further analysis; monovette 1 from each participant was immediately sent to the laboratories of the Department of Biological Sciences of UL for analysis, while monovettes 2-5 were kept in the transportation box and sent to the Haematology and Biochemistry Laboratory of University Hospital Limerick for analysis immediately after the 5th and final blood draw.

Following the initial baseline blood draw at approximately 09.05, the participant was fed a standardized breakfast, namely porridge, which was made with boiling water as per the manufacturer’s instructions (Flahavan’s Quick Oats® original with the following nutritional value: Energy 637 kJ/ 150 kcal; Fat 2.6 g; of which saturated 0.4 g; Carbohydrates 25 g; of which sugars 7.3 g; Fibre 3.5 g; Protein 5.2 g; Salt 0.03 g).

Once breakfast was consumed, randomized and double-blind administration of capsules of different types and dose, according to the aforementioned trials design took place. Each capsule was administered with a glass of water. The capsule compositions were as follows:

1. LDSNR, low-dose stomach non-resistant capsule containing 0.25 g of SPL
2. HDSNR, high-dose stomach non-resistant capsule contained 0.5 g of SPL
3. LDSR, low-dose stomach resistant capsule contained 0.25 g of SPL
4. HDSR, high-dose stomach resistant capsule contained 0.5 g of SPL
5. Placebo stomach release capsule containing 0.5 g of food-grade glycerin

Blood samples were subsequently obtained by repeating the steps for blood sampling every h for the next 4 h. The citrated blood samples were sent at each time of blood withdrawal to the Department of Biological Sciences of UL for platelet aggregometry testing and the remaining blood tubes were stored in a blood transportation box to be taken to the haematology and Biochemistry laboratories of University Hospital Limerick, where several biological parameters were assessed. After each blood draw, the cannula was flushed with saline (0.9 % w/v). During the study, participants were encouraged to drink at least 50 mL of water following every blood draw and were encouraged to keep their cannulated arm supported, still, and warm, while remaining seated.

### 2.3. Ex vivo human platelet-rich plasma (hPRP) aggregation assays

The platelet aggregation response to PAF and thrombin was conducted as previously described with some modifications [34]. Briefly, standard solutions of PAF and thrombin were prepared ranging from 2.6 x 10^-8^ to 2.6 x 10^-5^ mol/L BSA (2.5 mg BSA/mL saline (0.9%)) PAF and 0.01-0.4 U/mL active thrombin in saline (0.9%). hPRP was isolated from anticoagulated (sodium citrate 0.106 mol/l) blood samples by centrifugation at 194 x g for 18 min at 24 °C, the supernatant hPRP transferred to polypropylene tubes at room temperature. Platelet-poor plasma (PPP) was obtained by further centrifugation at 1465 x g for 20 min at 24°C and supernatant PPP transferred to polypropylene tubes at room temperature for aggregation bioassay. Then an aggregation bioassay was conducted. Briefly, PPP (250 μL) and hPRP (250 μL) were pipetted into separate glass aggregometer (Chronology, Havertown, PA, USA) cuvettes at 37 °C. Platelet aggregation, detected by change in light transmission at a fixed wavelength (620 nm) in the presence of increasing concentration of agonist generated aggregation curves from which the concentration of agonist that induces maximum-reversible platelet aggregation could be determined. The serial reduction of either PAF or thrombin required to induce maximum-reversible platelet aggregation revealed a linear response between 20%-80% of maximum and the concentration of agonist that induced 50% platelet aggregation, i.e., the EC_50_ value. The EC_50_ value represented platelet sensitivity to the agonist. The higher the EC_50_ value the lower the sensitivity of the platelet to the agonist to either PAF or thrombin.

### 2.2. Biochemical and haematological measuements

The University Hospital Limerick (Limerick, Ireland) undertook all biochemical and haematological analyses. Chemiluminescent magnetic microparticle immunoassay (CMIA) technology (Abbott Architect C16000 clinical chemistry analyser®, Abbott Ltd., Dublin, Ireland) was employed to measure glucose, triglycerides (TG), total cholesterol (TC), high-density lipoprotein cholesterol (HDL-C), low-density lipoprotein cholesterol (LDL-C) and C-reactive protein (CRP) in each blood sample. Prothrombin time (PT) activated partial thromboplastin time (APTT), fibrinogen and the international normalized ratio (INR) were measured by electromechanical detection (Stago STA-R Evolution® analyser, Diagnostica Stago, Wicklow, Ireland).

### 2.2. Statistical analysis

This pilot study employed a randomized, double-blind, placebo-controlled crossover design with five conditions and repeated sampling (baseline plus four hourly draws). The a priori power calculation indicating ∼83% power to detect a 1-SD difference assumed a large treatment effect and favorable assumptions regarding between- and within-subject variability. Because repeated measures collected over a short time window are often highly correlated, the effective statistical power in this small sample (n = 5 completing all conditions) is likely lower than the nominal estimate. Accordingly, this study was primarily intended as a pilot to assess safety, tolerability, and variance characteristics rather than to provide definitive evidence of efficacy. All data are presented as a mean percentage EC_50_ with standard error mean. The percentage change in EC_50_ from baseline and the temporal percentage incremental change ((ΔCt-0/C0)) in EC_50_ from baseline were calculated for each time point for each treatment, which are depicted in Figures 2 and 3.

**Fig. 2.**
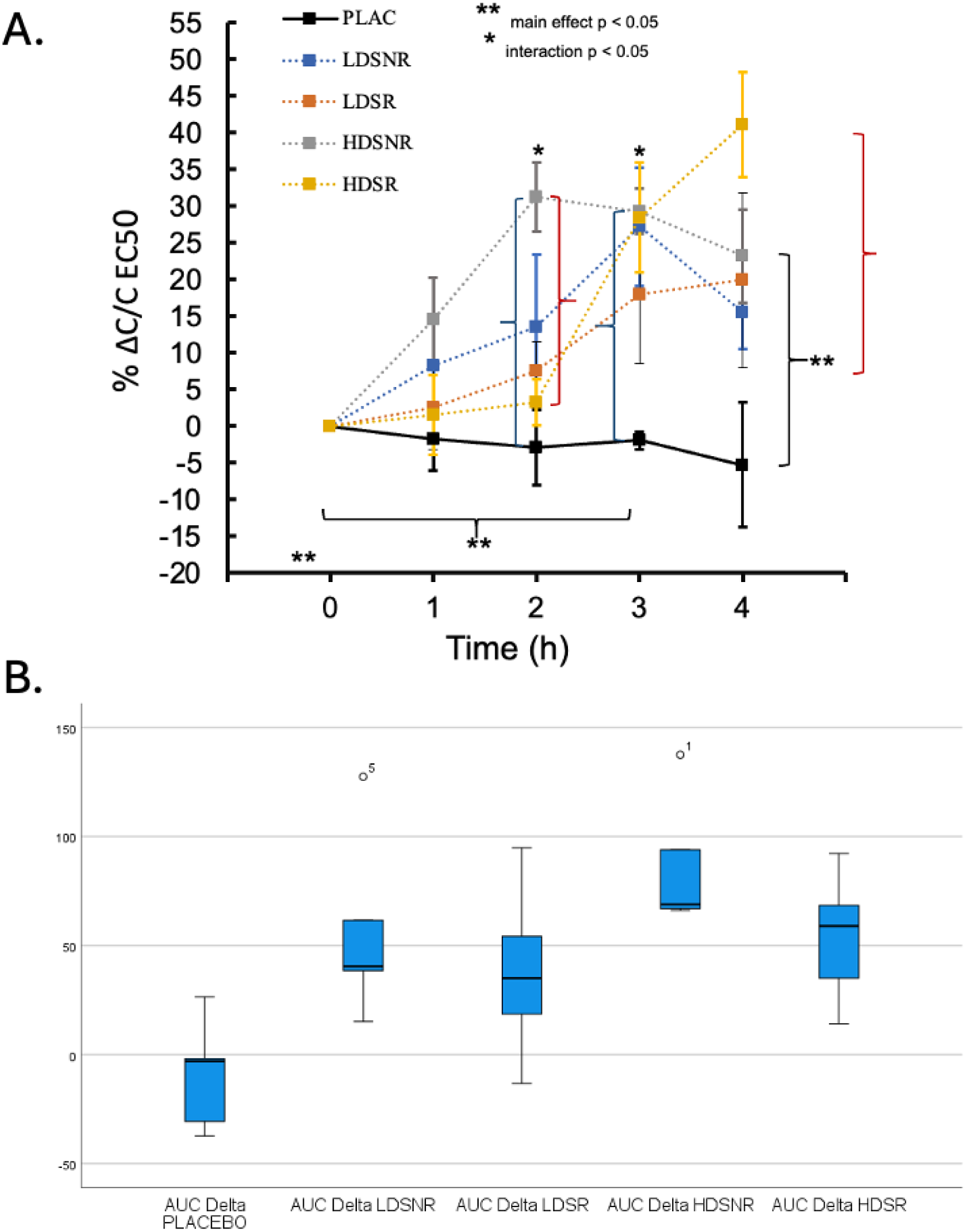
**(A)** Temporal change in EC_50_ values of PAF-induced platelet aggregation in human platelet-rich plasma across all treatment conditions. Data are presented as mean percentage change from baseline (ΔC/C EC_50_; n = 5) with error bars representing the standard error of the mean (SEM). Statistical analysis was performed using a two-factor repeated-measures ANOVA (treatment × time). Where significant interactions were detected, Bonferroni-adjusted pairwise comparisons were applied. *Indicates *p* < 0.05 for significant pairwise differences between treatments at a given time point. **Indicates *p* < 0.05 for significant main effects of treatment or time. An increase in EC_50_ reflects a reduction in platelet sensitivity to PAF. **(B)** Boxplots showing the area-under-the-curve (AUC) for ΔPAF EC_50_ (0–4 h) across all treatment conditions. Boxes represent the interquartile range (IQR), horizontal lines indicate the median, and whiskers show the range excluding outliers; circles represent values >1.5 × IQR identified during outlier screening. AUC data were analysed using a repeated-measures ANOVA after confirming normality (Shapiro–Wilk) and sphericity (Mauchly’s test). A significant main effect of treatment was detected (*p* < 0.05), with post-hoc contrasts demonstrating higher ΔAUC in the HDSNR condition compared with placebo. **Abbreviations:** HDSNR, high-dose stomach non-resistant capsule contained 0.5 g of SPL; IQR, interquartile range; HDSR, high-dose stomach resistant capsule contained 0.5 g of SPL; LDSNR, low-dose stomach non-resistant capsule containing 0.25 g of SPL; LDSR, low-dose stomach resistant capsule contained 0.25 g of SPL. PAF, platelet-activating factor; PLAC, placebo control.

**Fig. 3.**
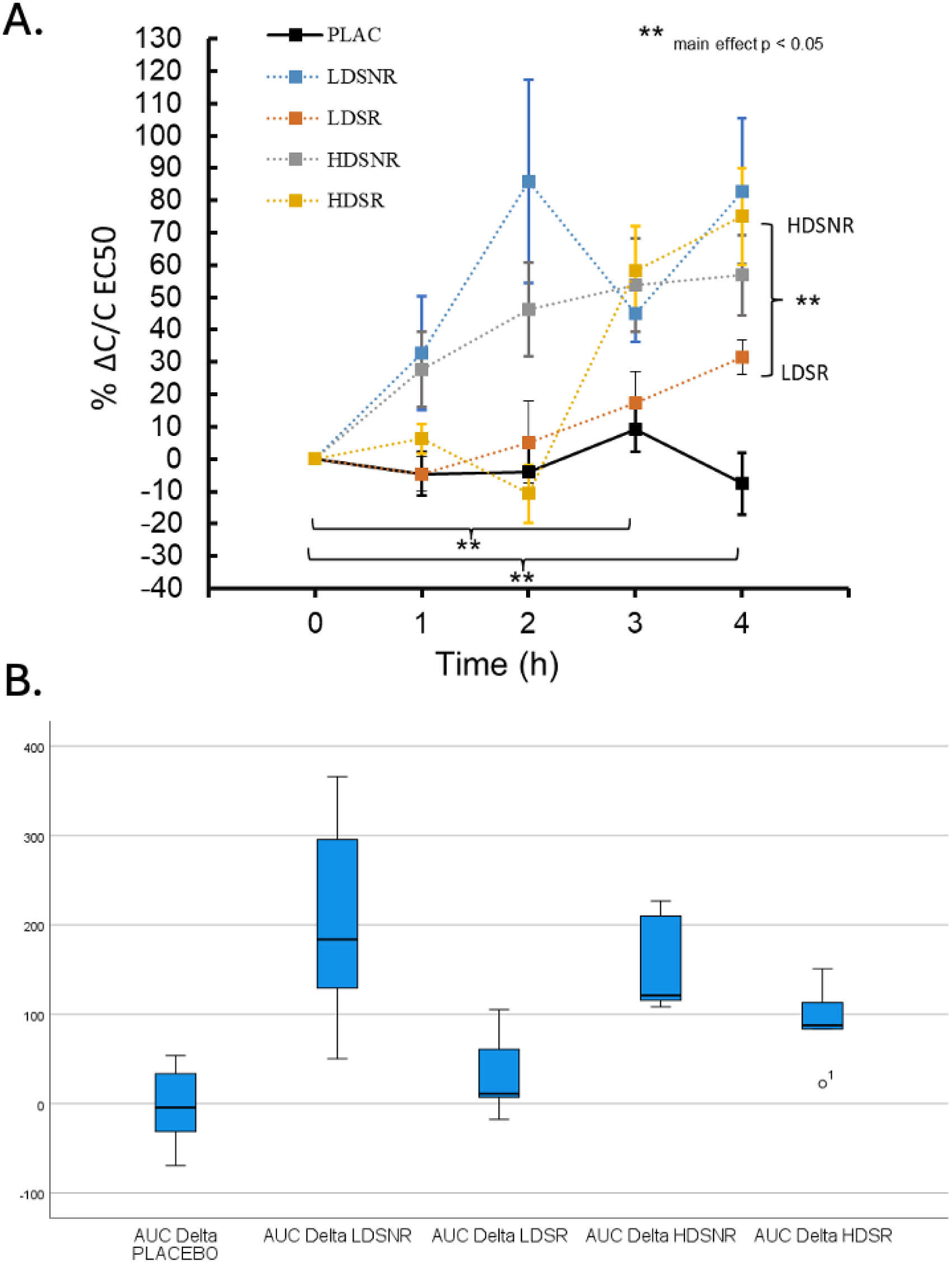
**(A)** Temporal change in EC_50_ values of thrombin-induced platelet aggregation of human platelet-rich plasma across all treatment conditions. Data are presented as mean percentage change from baseline (ΔC/C EC_50_; n = 5) with error bars representing the standard error of the mean (SEM). Statistical analysis was performed using a two-factor repeated-measures ANOVA (treatment × time). No significant interaction was detected; however, significant main effects of treatment and time were observed, with post-hoc pairwise comparisons applied where appropriate. Higher EC_50_ values indicate reduced platelet sensitivity to thrombin. **(B)** Boxplots showing the area-under-the-curve (AUC) for Δthrombin EC_50_ (0–4 h) for each treatment. Boxes represent the interquartile range (IQR), horizontal lines indicate the median, and whiskers show the range excluding outliers; circles denote values >1.5 × IQR identified during outlier screening. AUC data were analysed using repeated-measures ANOVA, which revealed a significant main effect of treatment. Post-hoc pairwise comparisons indicated that HDSNR elicited a greater cumulative change in thrombin EC50 compared with LDSR. **Abbreviations:** HDSNR, high-dose stomach non-resistant capsule contained 0.5 g of SPL; IQR, interquartile range; HDSR, high-dose stomach resistant capsule contained 0.5 g of SPL; LDSNR, low-dose stomach non-resistant capsule containing 0.25 g of SPL; LDSR, low-dose stomach resistant capsule contained 0.25 g of SPL. PAF, platelet-activating factor; PLAC, placebo control.

For analysis of the PAF EC_50_ data, there were no outliers as shown by examination of studentized residuals for values greater than ± 3. The data was assessed for normality distributed (*p >* 0.05) apart from 1h LDSNR and 3h of HDSNR (*p* < 0.05) as assessed by Shapiro-Wilk’s test. Mauchly’s test of sphericity indicated that the assumption of sphericity was not met for the two-way interaction, χ^2^(2) = 67.6, *p* = 0.003 and Greenhouse-Geisser correction applied. A two-factor analysis of variance (ANOVA; treatment x time) was used with repeated measures on the factor time. The Bonferroni post hoc correction was applied where significant interaction was evident.

For PAF ΔAUC, outliers were assessed via boxplots (values > 1.5 box-lengths). Shapiro–Wilk’s test indicated normal distribution for all treatment conditions except HDSNR, which showed marginal deviation (*p* = 0.048). Mauchly’s test indicated that the assumption of sphericity was met; therefore, no correction was applied (χ^2^(9) = 12.52, *p* = 0.281). Significance was set at ***p*** < 0.05, and effect sizes are reported as partial eta squared (η_p_²). Post-hoc comparisons were conducted using pairwise contrasts with adjusted *p-*values where appropriate. All EC_50_ outcomes are presented as percentage change from baseline (mean ± SEM). Statistical results in text are reported as mean differences with 95% confidence intervals (CI)

For thrombin EC_50_, data screening identified no outliers based on studentized residuals (±3). Normality and sphericity were assessed as described for PAF; sphericity was violated for the treatment × time interaction (χ^2^(2) = 67.6, *p* = 0.003), and Greenhouse–Geisser correction was applied. Thrombin ΔAUC analysis identified one outlier in the HDSR condition using the boxplot criterion (>1.5 × IQR). AUC data met normality assumptions, and sphericity was not violated; therefore, no correction was required. Repeated-measures ANOVA was used for both EC_50_ and AUC analyses, with post-hoc pairwise comparisons performed where appropriate. All analyses were conducted using Microsoft Excel (v16.1.1) and SPSS (v30.0.0).

## 3. Results

### 3.1. Postprandial effect of SPL on plasma levels of CRP, glucose, and lipid levels

Baseline plasma biomarkers of the lipid profile TG, TC, HDL-C, LDL-C, glucose, and CRP were within the normal range prior to the study for all participants (Table S1) and no significant change in these biomarkers was observed following ingestion of the SPL or placebo capsules within the timeframe assessed (Figure S1). Cumulative ingestion and exposure should be assessed in a dietary study.

### 3.2. Postprandial effect of SPL on biomarkers of coagulation

Baseline plasma fibrinogen, PT, INR, and APTT were within normal range for all participants prior to the study (Table S2). Furthermore, these parameters did not differ significantly from baseline following ingestion of the SPL or placebo capsules within the timeframe assessed but further study is required for cumalitve exposure (Figure S2).

### 3.3. Postprandial effect of SPL on platelte sensitivity to PAF

Figure 2A depicts the temporal change in PAF EC_50_ for each treatment following ingestion when compared to baseline levels (t = 0). The placebo did not significantly affect PAF EC_50_ at any time during the 4 h postprandial period.

Analysis of ΔPAF EC_50_ revealed a significant treatment × time interaction, indicating that the temporal pattern of change differed across treatments (F(16, 64) = 3.30, *p* ≤ .047, η_p_² = 0.452). Post-hoc pairwise comparisons showed that HDSNR produced significantly greater increases in PAF EC_50_ compared with both placebo and HDSR at specific time points. At 2 h, ΔPAF EC_50_ was higher in HDSNR versus placebo (mean difference 34.16, 95% CI 4.67 to 63.65, *p* = .029) and versus HDSR (27.9, 95% CI 1.9 to 53.9, *p* = .039). At 3 h, ΔPAF EC_50_ remained elevated relative to placebo (31.2, 95% CI 7.9 to 54.6, *p* = .017). Within-condition comparisons showed that HDSNR elicited a significant rise from baseline at both 2 h (31.2, 95% CI 4.89 to 57.5, *p* = .027) and 3 h (29.3, 95% CI 11.9 to 46.7, *p* = .007). For the HDSR condition, increases were only evident at 4 h (e.g., 41.1 above baseline, 95% CI 1.17 to 80.9, *p* = .045).

When collapsed across time, a significant main effect of treatment was also observed (F(4,16) = 6.75, *p* = .015, η_p_² = 0.628), driven by a higher mean ΔPAF EC_50_ in HDSNR compared with placebo (22.04, 95% CI 13.7 to 30.4, *p* = .001). A significant main effect of time indicated that ΔPAF EC_50_ rose above baseline at 3 h across treatments (20.16, 95% CI 3.19 to 37.2, *p* = .027). For ΔAUC (Figure 2B), repeated-measures ANOVA identified a significant treatment effect (F(4, 16) = 5.784, *p* = .004, η_p_² = 0.591). Post-hoc pairwise comparisons showed that HDSNR produced a greater cumulative change in EC_50_ over 0–4 h compared with placebo (mean difference 95.96, 95% CI 55.06–136.7, *p* = .002). No other pairwise comparisons were significant.

### 3.4. Postprandial effect of SPL on platelet sensitivity to thrombin

Figure 3 depicts the temporal change in thrombin EC_50_ for each treatment following ingestion when compared to baseline measurements (t = 0). The placebo did not significantly affect thrombin EC_50_ at any time in the 4 h postprandial period. Analysis of Δthrombin EC_50_ revealed no significant treatment × time interaction (F(16, 64) = 3.823, *p* = .050, η_p_² = 0.489), indicating that temporal changes were similar across treatments. When examined independently of time, there was a significant main effect of treatment (F(4, 16) = 8.965, *p* = .007, η_p_² = 0.691). Post-hoc pairwise comparisons showed that Δthrombin EC_50_ was significantly higher following HDSNR compared with LDSR (mean difference 27.1, 95% CI 10.6–43.6, *p* = .008). A significant main effect of time was also observed (F(4, 16) = 16.221, *p* < 0.001, η_p_² = 0.802). Compared with baseline, Δthrombin EC_50_ increased significantly at 3 h (36.7, 95% CI 10.8–62.6, *p* = .014) and 4 h (47.7, 95% CI 17.0–78.5, *p* = .010) across treatments. For the ΔAUC analysis, one mild outlier (HDSR) was detected (Figure 3). Repeated-measures ANOVA identified a significant effect of treatment on thrombin ΔAUC (F(4, 16) = 8.555, *p* < 0.001, η_p_² = 0.681). Post-hoc pairwise comparisons indicated that HDSNR produced a significantly greater cumulative increase in thrombin EC50 over 0–4 h compared with LDSR (mean difference 122.9, 95% CI 53.9–191.9, *p* = .006). No additional pairwise differences reached significance.

## 4. Discussion

Activation and aggregation of platelets contribute to the inflammatory and thrombotic manifestations of atherosclerosis and CVD [5,6,12]. Dysregulated platelet activity also plays an important role in other pathologies, including type II diabetes mellitus [35] and cancer [36]. Repetitive activation of platelets and plasmatic coagulation during the postprandial period has further been implicated in the initiation, development, and progression of atherosclerosis [2,21,37]. These mechanisms highlight the importance of identifying bioactive molecules and natural products capable of attenuating platelet activation, with the aim of developing novel supplements and nutraceuticals with potential cardioprotective properties. While there are several avenues of potential research to explore, including dietary components of the Mediterranean diet capable of reducing postprandial platelet sensitivity to PAF in healthy individuals and patients with metabolic syndrome [38,39].

Marine-derived polar lipids represent one such promising class of bioactives. Previous in vitro studies have demonstrated that salmon polar lipid (SPL) extracts contain bioactive molecules with anti-PAF and anti-thrombin activities [25,27,28]. A broader body of work from our group and others has shown that marine polar lipids exhibit diverse anti-inflammatory and cardioprotective effects [17,23,27,31,40,41], including anti-atherogenic actions in platelets and various cell types both *in vitro* and *in vivo* [42,43]. While natural antiplatelet nutraceuticals such as FruitFlow™, a water soluble (lipid-free) tomato-based supplement shown to beneficially affect postprandial ADP-induced platelet aggregation [44], have reached commercial use, targeted clinical work examining marine polar lipid preparations remains limited.

In this context, the present pilot study provides the first in vivo indication that SPL ingestion may modestly reduce platelet sensitivity to both PAF and thrombin in healthy subjects. Among the five postprandial trials tested, the high-dose non–stomach-resistant capsule (HDSNR) produced the clearest reductions in platelet sensitivity, with significant increases in EC_50_ for PAF at 2–3 h and for thrombin at 3–4 h post-ingestion. The stomach-resistant formulation (HDSR) generated similar but delayed effects, suggesting that the timing of polar lipid release plays a crucial role in determining the onset of antiplatelet activity. The low-dose capsules produced more modest or inconsistent responses. Despite the small sample size, these findings support the concept that SPL-derived polar lipids acutely modulate platelet reactivity, potentially through interactions with the PAF-R and PAR receptors. The stronger and earlier effect observed with HDSNR is consistent with more rapid release and absorption of bioactive lipids, whereas the attenuated and delayed effect of HDSR suggests slower release in the gastrointestinal tract.

Importantly, these antiplatelet effects occurred without measurable alterations in key haemostatic biomarkers (fibrinogen, PT, APTT, INR). Monitoring such biomarkers is essential when evaluating novel antiplatelet agents to ensure that modulation of platelet sensitivity does not translate into increased bleeding risk [45,46]. Notably, the reduction of the sensitivity of platelets against both PAF and thrombin during both high-dose trials does not seem to affect coagulation biomarker (Fibrinogen, PT, APTT, and INR). These results further suggest that the observed favorable postprandial antiplatelet effects against both PAF and thrombin do not postprandially increase the risk of bleeding, even when supplements containing high doses of the SPL were administrated during the HDSNR and HDSR trials. These findings support the notion that marine phospholipid supplements are safe for consumption [31]. However, cumulative intake and long-term exposure must be monitored by conducting a dietary intervention study to ensure safety.

Plasma fibrinogen, an acute-phase reactant associated with cardiovascular risk and platelet aggregation [46], also remained unchanged. The absence of fibrinogen fluctuations likely reflects the healthy status of the cohort, as supported by consistently low CRP values (<5 mg/L) throughout the study. Similarly, postprandial glucose and lipid levels (TC, HDL-C, LDL-C, TG) remained stable, which is desirable given that postprandial hyperglycemia and lipemia can augment platelet reactivity and cardiovascular risk [20,47,48]. The stability of these markers likely reflects the healthy metabolic profile of participants at baseline.

The earlier and more pronounced antiplatelet response observed in the HDSNR trial compared with the HDSR trial likely reflects differences in capsule disintegration and bioactive lipid release. Non-resistant capsules permit immediate release in the stomach, whereas resistant capsules delay release until later digestive stages. These release profiles may have implications for optimizing postprandial timing and therapeutic efficacy with further considerations for circadian rhythms [49,50]. Nonetheless, the overall magnitude of the antiplatelet response was modest and must be interpreted with caution, as intra- and inter-individual variability may have influenced outcomes in this small cohort. Platelet function is known to vary with diet, ethnicity, sex, and other factors [51]. Furthermore, a postprandial assessment is required to determine how long the postprandial effects lasts, to inform for dosage in further studies.

While consumption of the capsule in this trial appears to be safe and did not increase the risk of bleeding following acute exposure, further long-term studies are required to confirm these observations. Furthermore, we did not observe how long the antiplatelet effect would last postprandially. Therefore, additional research is required to determine whether a repeated dose schedule is required. Previous studies of omega-3 fatty acid preparations, which carry similar fatty acids but in neutral forms, have shown that the antiplatelet effects may last up to hours post ingestion. However, it is difficult to determine whether this would be a similar response using PL supplements and our findings would suggest that the encapsulation method would be important to consider.

Additionally, this study used a relatively low (0.25 g) and high-dose (0.5 g) version of the capsule, similar trials using fish oils high in EPA and DHA known to have antithrombotic effects previously administered doses in excess of 5g [52]. A conservative amount of fish oil in the form of polar lipids used was chosen for these trials due to the greater reported bioavailability and bioactivity of polar lipids. Future work should evaluate higher SPL doses (e.g., 1 g) to better characterize dose–response relationships. Tolerability was good in this study, with no gastrointestinal symptoms or vasoconstrictive adverse events reported. However, side effects are possible and fish oil supplements have been associated with diarrhea, but these side effects tend to be associated with large doses of fish oil [52]. Fishy breath or belching is also a possible side effect of fish oil supplement intake. Indeed, the salmon capsules used in this study were not deodorized, as is common for similar fish supplements. We chose not to undertake this step due to potential degradation of the lipid content [53]. Therefore, dietary intervention studies should assess for these common side effects to improve formulation of the capsule.

Lastly, the diet of fish influences the fatty acid and polar lipid composition of the flesh, which in turn affects the composition of fish oils [54,55]. This opens the field to opportunities for tailored polar lipid compositions as a therapeutic or as a liposomal carrier [56] for other drugs through modulation of the diet in aquaculture setting.

## 5. Conclusions

This is the first placebo-controlled study reporting a modest favorable outcome of the administration of a novel food supplement containing bioactive SPL, with respect to postprandial platelet sensitivity against PAF and thrombin and its related cardiovascular risk in healthy human subjects. This pilot study provides data and the rationale to support the development of further large long-term trials to confirm these findings in both healthy subjects and cardiovascular patients to evaluate the long-term effects, safety, tolerability, and cardioprotective properties of this novel supplement.

## Supplementary Materials

Table S1: Participant baseline physical and haematological characteristics; Table S2: Participant baseline biochemical characteristics. Figure S1: Postprandial effect of SPL on the plasma levels of triglycerides, total cholesterol, HDL, and LDL cholesterol and glucose; Figure S2: Postprandial effect of SPL on blood coagulation biomarkers.

## Author Contributions

Conceptualization, I.Z., A.T., P.J. and R.L.; methodology, R.L. and A.T.; software, R.L.; validation, P.J., I.Z. and R.L.; formal analysis, A.T. and R.L.; investigation, A.T. and R.L..; resources, P.J.I.Z.; data and curation, R.L. and P.J.; writing—original draft preparation, R.L. and A.T.; writing—review and editing, R.L., A.T., P.J. and I.Z.; visualization, A.T., R.L. and P.J.; supervision, P.J. and I.Z.; project administration. A.T. and R.L.; funding acquisition, P.J., R.L. and I.Z. All authors have read and agreed to the published version of the manuscript.

## Institutional Review Board Statement

The study was conducted in accordance with the Declaration of Helsinki and approved by the Faculty of Science & Engineering Research Ethics Committee of the University of Limerick (UL Ethics Approval Number: 2017_10_10_S&E).

## Informed Consent Statement

Informed consent was obtained from all subjects involved in the study.

## Data Availability Statement

Data is available from the authors without reservation.

## Acknowledgments

The authors are grateful to the volunteers who took part in the study, to Elaine Ahern for her phlebotomy support, and to Dr Emmet Kerrin at Treaty Medical, Limerick. The authors acknowledge the support of Lifes2good, Marine Harvest, and Mowi Ireland for their contributions. We would also like to thank the Departments of Physical Education and Sports Science and the Biological Sciences at the University of Limerick, Ireland, for their continued support. Finally, we are grateful to Patricia Kennedy, Carolyn Holt, and colleagues at the laboratories of the University Hospital Limerick for analytical support.

