## Supplementary File for "Encapsulated Salmon Polar Lipids Modestly Reduce Postprandial Platelet Sensitivity to PAF and Thrombin in Healthy Adults: A Pilot Study"

**Supplementary Materials:**

**Table S1:** Participant baseline physical and haematological characteristics.

| | **Participants (n = 5)** |
|---|---|
| Female | n = 3 |
| Male | n = 2 |
| Age | 33.8 (28-48) |
| BMI (kg/m$^2$) | 24.9 (18.8-29.7) |
| Resting heart rate | 68 (57-74) |
| Blood pressure (mmHg) | |
| Systolic | 110 (100-120) |
| Diastolic | 69 (57-80) |
| WBC ($10^9$/L) | 6.3 (5.5-7.4) |
| RBC ($10^9$/L) | 4.8 (4.4-5.4) |
| Platelets ($10^9$/L) | 265 (161-410) |
| Hb (g/dL) | 13.6 (12-15) |
| Hematocrit (%) | 42.7 (40.1-47.2) |

BMI = Body mass index; Hb = Haemoglobin; RBC = Red blood cells; WBC = White blood cells

**Table S2:** Participant baseline biochemical characteristics.

| | Participants (n = 5) |
|---|---|
| Plasma lipids (mmol/L) | |
| Total Cholesterol | 4.2 (3.4-5.2) |
| LDL Cholesterol | 2.6 (2.0-3.1) |
| HDL Cholesterol | 1.5 (1.2-2.0) |
| Triglycerides | 0.7 (0.5-1.0) |
| Other Plasma Constituents | |
| Glucose (mmol/L) | 4.6 (4.3-5.0) |
| hsCRP (mg/L) | < 5 |
| Ferritin (ng/mL) | 51.4 (11.0-98.0) |
| HbA1c (mmol/mol) | 34.6 (32.0-38.0) |

HbA1c = Hemoglobin A1c; HDL = high-density lipoprotein; hsCRP = high-sensitivity C-reactive protein; LDL = low-density lipoprotein

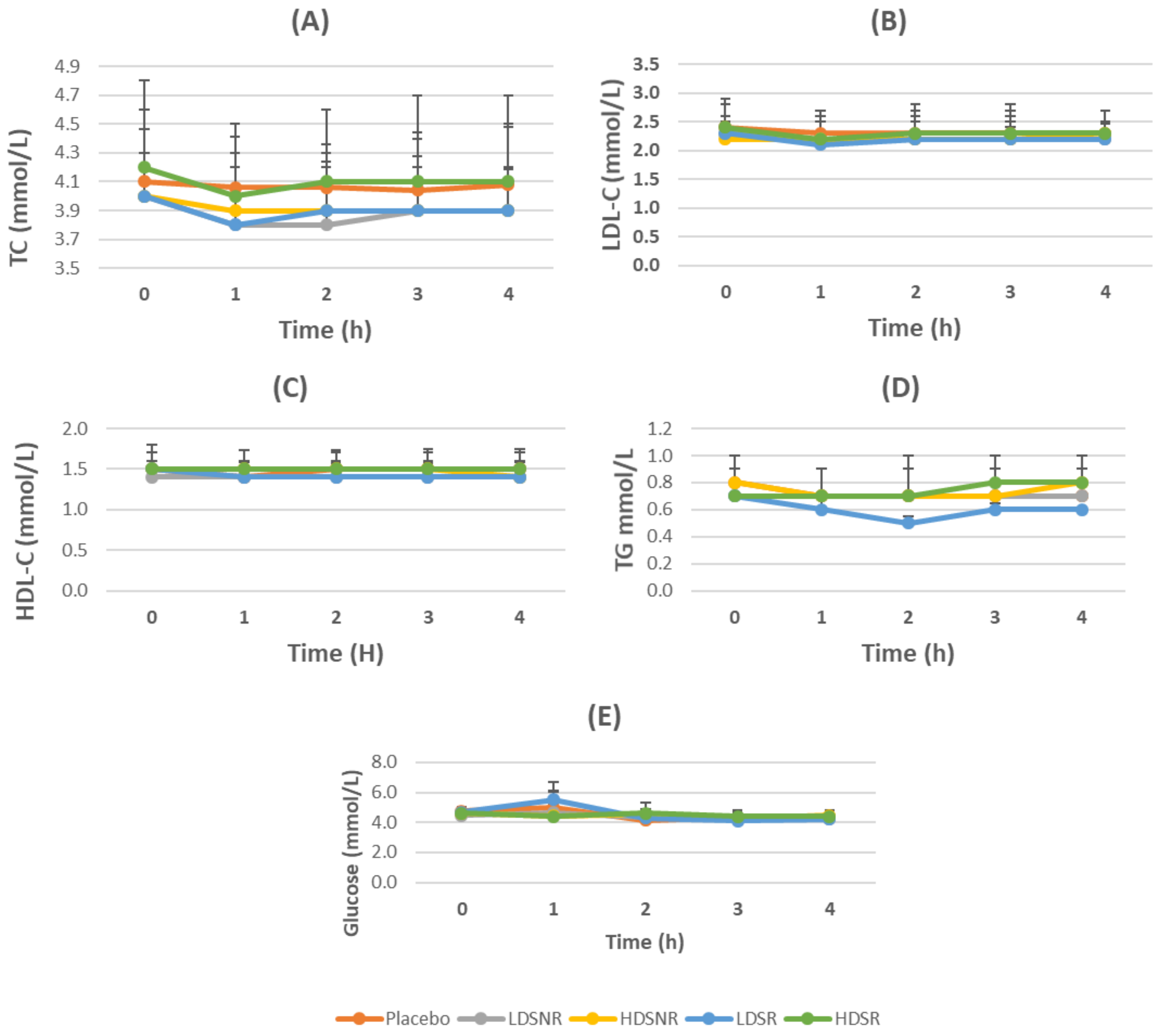


**Figure S1.** Postprandial effect of FGSPL on the plasma levels of triglycerides, total cholesterol, HDL, and LDL cholesterol and glucose. Data are the mean (n = 5). Error bars represent the standard deviation..

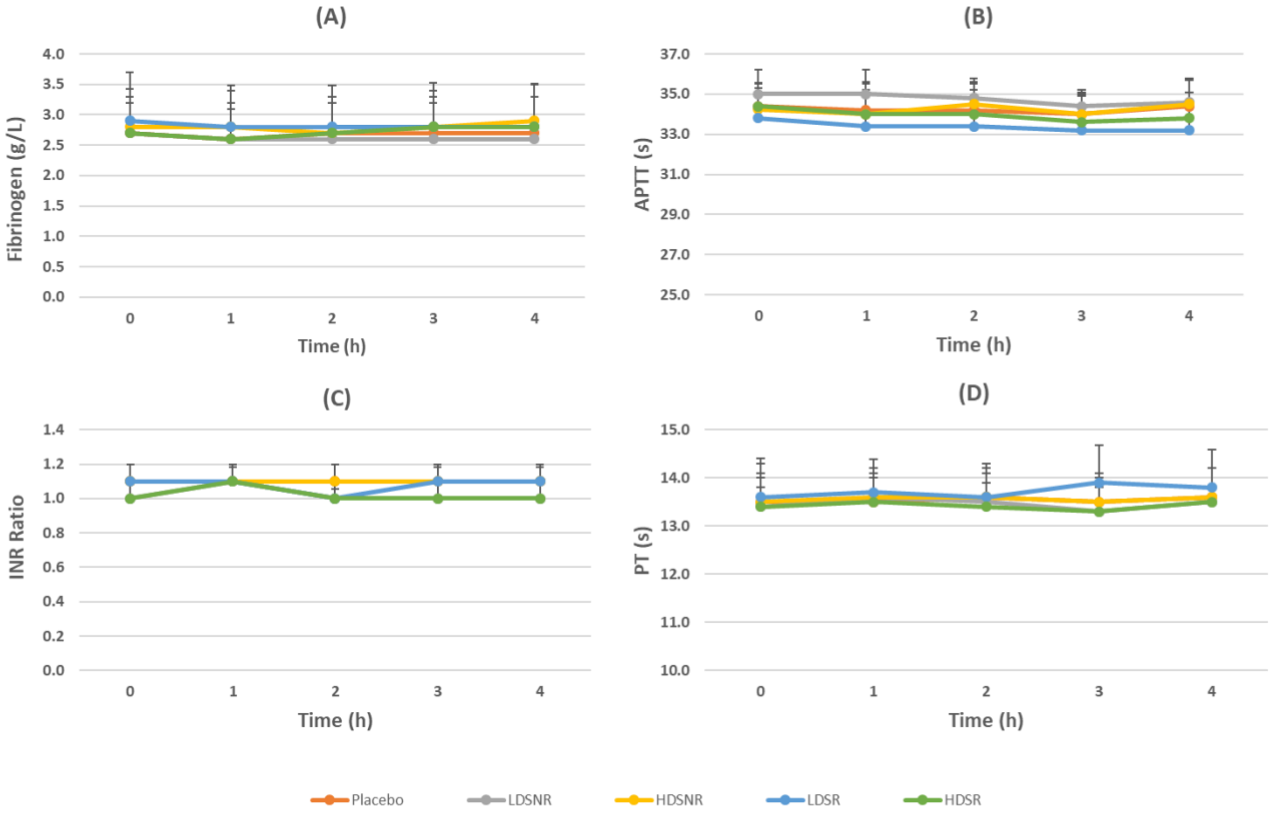


**Figure S2.** Postprandial effect of FGSPL on blood coagulation biomarkers: Fibrinogen (g/L), prothrombin time (PT, s), international normalised ratio (INR), activated partial thromboplastin time (APTT, s). Data are the mean (n = 5). Error bars represent the standard deviation.
